# OpenSAFELY: factors associated with COVID-19-related hospital death in the linked electronic health records of 17 million adult NHS patients

**DOI:** 10.1101/2020.05.06.20092999

**Authors:** The OpenSAFELY Collaborative, Elizabeth Williamson, Alex J Walker, Krishnan Bhaskaran, Seb Bacon, Chris Bates, Caroline E Morton, Helen J Curtis, Amir Mehrkar, David Evans, Peter Inglesby, Jonathan Cockburn, Helen I McDonald, Brian MacKenna, Laurie Tomlinson, Ian J Douglas, Christopher T Rentsch, Rohini Mathur, Angel Wong, Richard Grieve, David Harrison, Harriet Forbes, Anna Schultze, Richard Croker, John Parry, Frank Hester, Sam Harper, Raf Perera, Stephen Evans, Liam Smeeth, Ben Goldacre

**Affiliations:** The DataLab, Nuffield Dept of Primary Care Health Sciences, University of Oxford, OX2 6GG; London School of Hygiene and Tropical Medicine, Keppel Street, London WC1E 7HT; TPP, TPP House, 129 Low Lane, Horsforth, Leeds, LS18 5PX; ICNARC, 24 High Holborn, Holborn, London WC1V 6AZ; NIHR Health Protection Research Unit (HPRU) in Immunisation

**Keywords:** COVID-19, risk factors, ethnicity, deprivation, death, informatics

## Abstract

**Background:** Establishing who is at risk from a novel rapidly arising cause of death, and why, requires a new approach to epidemiological research with very large datasets and timely data. Working on behalf of NHS England we therefore set out to deliver a secure and pseudonymised analytics platform inside the data centre of a major primary care electronic health records vendor establishing coverage across detailed primary care records for a substantial proportion of all patients in England. The following results are preliminary.

**Data sources:** Primary care electronic health records managed by the electronic health record vendor TPP, pseudonymously linked to patient-level data from the COVID-19 Patient Notification System (CPNS) for death of hospital inpatients with confirmed COVID-19, using the new OpenSAFELY platform.

**Population:** 17,425,445 adults.

**Time period:** 1st Feb 2020 to 25th April 2020.

**Primary outcome:** Death in hospital among people with confirmed COVID-19.

**Methods:** Cohort study analysed by Cox-regression to generate hazard ratios: age and sex adjusted, and multiply adjusted for co-variates selected prospectively on the basis of clinical interest and prior findings.

**Results:** There were 5683 deaths attributed to COVID-19. In summary after full adjustment, death from COVID-19 was strongly associated with: being male (hazard ratio 1.99, 95%CI 1.88-2.10); older age and deprivation (both with a strong gradient); uncontrolled diabetes (HR 2.36 95% CI 2.18-2.56); severe asthma (HR 1.25 CI 1.08-1.44); and various other prior medical conditions. Compared to people with ethnicity recorded as white, black people were at higher risk of death, with only partial attenuation in hazard ratios from the fully adjusted model (age-sex adjusted HR 2.17 95% CI 1.84-2.57; fully adjusted HR 1.71 95% CI 1.44-2.02); with similar findings for Asian people (age-sex adjusted HR 1.95 95% CI 1.73-2.18; fully adjusted HR 1.62 95% CI 1.43-1.82).

**Conclusions:** We have quantified a range of clinical risk factors for death from COVID-19, some of which were not previously well characterised, in the largest cohort study conducted by any country to date. People from Asian and black groups are at markedly increased risk of in-hospital death from COVID-19, and contrary to some prior speculation this is only partially attributable to pre-existing clinical risk factors or deprivation; further research into the drivers of this association is therefore urgently required. Deprivation is also a major risk factor with, again, little of the excess risk explained by co-morbidity or other risk factors. The findings for clinical risk factors are concordant with policies in the UK for protecting those at highest risk. Our OpenSAFELY platform is rapidly adding further NHS patients’ records; we will update and extend these results regularly.

## Introduction

On March 11th 2020, the World Health Organisation characterised COVID-19 as a pandemic after 118,000 cases and 4,291 deaths were reported in 114 countries.^1^ As of 30 April, cases are over 3 million globally, with more than 200,000 deaths attributed to the virus.^2^ In the UK, cases have reached 171,253, with 22,791 deaths in hospital.^3^

Age and gender are well-established risk factors, with over 90% of UK deaths to date being in people aged over 60 years, and 60% of deaths in men,^4^ consistent with global patterns. Various pre-existing conditions have been reported to correlate with increased risk of poor outcomes. In a re-analysis of a large aggregated case series dataset from the Chinese center for disease control and prevention (44,672 patients, 1,023 deaths), cardiovascular disease, hypertension, diabetes, respiratory disease, and cancers were all associated with increased risk of death.^5^ These factors often correlate with age, but correction for age was not possible in the available data. More recently, a large UK cross-sectional survey describing 16,749 patients already hospitalised with COVID-19 showed higher risk of death for patients with cardiac, pulmonary and kidney disease, as well as malignancy, dementia and obesity (hazard ratios 1.19-1.39 after age and sex correction).^6^ Obesity has been reported as a risk factor for treatment escalation in a French ITU cohort (n=124) and a New York hospital presentation cohort (n=3615).^7,8^ The risks associated with smoking are disputed: increased risks were initially reported; recent studies suggest that smokers are underrepresented among those with more severe disease; and a potential protective mechanism for nicotine has been suggested.^9^ Smoking prevalence among hospitalised patients was lower than expected in China (1,099 patients, 12.6% vs 28% in the general population),^10^ and in a small French study (139 outpatients and 343 inpatients; Standardized Incidence Ratios 0.197 and 0.246, respectively).^11^ People from black and minority ethnic (BME) groups are at increased risk of bad outcomes from COVID-19, but explanations for this association are unclear.^12,13^

We therefore set out to determine factors associated with risk of death from COVID-19 in England using a very large sample of the adult population, with deaths data linked to longitudinal primary care electronic health records. This is the first iteration, based on the currently available data; further updates and additional outcomes will be released as more data become available through the OpenSAFELY.org platform.

## Methods

### Study design

We conducted a cohort study using national primary care electronic health record data linked to in-hospital COVID-19 death data (see Data Source). The cohort study began on 1st February 2020, chosen as a date several weeks prior to the first reported COVID-19 deaths and the day after the second laboratory confirmed case;^14^ and ended on 25th April 2020. The cohort explores risk among the general population rather than in a population infected with SARS-COV-2. Therefore, all patients were included irrespective of their SARS-COV-2 test results.

### Data Source

We used patient data from general practice (GP) records managed by the GP software provider The Phoenix Partnership (TPP), linked to COVID-19 inpatient hospital death notifications (CPNS) from NHSE/X, and Office for National Statistics (ONS) death data. CPNS provides the most current information on deaths of inpatients with laboratory confirmed COVID-19 occurring within NHS hospitals;^15,16^ whereas ONS includes information on all deaths, including those due to non-COVID-19 causes, and was used for censoring.

The data were accessed, linked and analysed using OpenSAFELY, a new data analytics platform created to address urgent questions relating to the epidemiology and treatment of COVID-19 in England. OpenSAFELY provides a secure software interface that allows detailed pseudonymised primary care patient records to be analysed in near realtime where they already reside, hosted within the EHR vendor’s highly secure data centre, to minimise the re-identification risks when data are transported off-site; other smaller datasets are linked to these data within the same environment using a matching pseudonym derived from the NHS number. More information can be found on https://opensafely.org/.

The dataset analysed with OpenSAFELY for this paper is based on 24 million currently registered patients (approximately 40% of the English population) from GP surgeries using the TPP SystmOne electronic health record system. It extends to 20 billion rows of structured data characterising (for example) pseudonymised patients’ diagnoses, medications, physiological parameters, and prior investigations. Data extracted from TPP SystmOne have previously been used in medical research, as part of the ResearchOne dataset.^17,18^

### Study Population and Observation Period

Our study population consisted of all adults (males and females 18 years and above) currently registered as active patients in a TPP general practice in England on 1st February 2020. To be included in the study, participants were required to have at least 1 year of prior follow-up in the GP practice to ensure that baseline patient characteristics could be adequately captured, and to have a recorded sex and age.^19^ Patients were observed from the 1st of February 2020 and were followed until the first of either their death date (whether COVID-19 related or due to other causes) or the study end date, 25th April 2020. For this analysis, CPNS death data were available up to 25th April 2020; ONS death data (used for censoring individuals who died without the outcome) were available to 16th April 2020; patient censoring for deaths due to other causes was therefore not possible during the last 9 days of followup (see Discussion, weaknesses; a sensitivity analysis is presented with all data censored at 6th April 2020 in appendix Table A1).

### Outcomes

The outcome was in-hospital death among people with confirmed COVID-19, ascertained from the COVID-19 Patient Notification System (CPNS).

### Covariates

Potential risk factors included: health conditions listed in UK guidance on “higher risk” groups;^20^ other common conditions which may cause immunodeficiency inherently or through medication (cancer and common autoimmune conditions); and emerging risk factors for severe outcomes among COVID-19 cases (such as raised blood pressure).

Age, sex, body mass index (BMI; kg/m^2^), and smoking status were considered as potential risk factors. Where categorised, age groups were: 18-<40, 40-<50, 50-<60, 60-<70, 70-<80, 80+ years. BMI was ascertained from weight measurements within the last 10 years, restricted to those taken when the patient was over 16 years old. Obesity was grouped using categories derived from the World Health Organisation classification of BMI: no evidence of obesity <30 kg/m^2^; obese I 30-34.9; obese II 35-39.9; obese III 40+. Smoking status was grouped into current, former and never smokers

The following comorbidities were also considered potential risk factors: asthma, other chronic respiratory disease, chronic heart disease, diabetes mellitus, chronic liver disease, chronic neurological diseases, common autoimmune diseases (Rheumatoid Arthritis (RA), Systemic Lupus Erythematosus (SLE) or psoriasis), solid organ transplant, asplenia, other immunosuppressive conditions, cancer, evidence of reduced kidney function, and raised blood pressure or a diagnosis of hypertension.

Disease groupings followed national guidance on risk of influenza infection,^21^ therefore “chronic respiratory disease (other than asthma)” included COPD, fibrosing lung disease, bronchiectasis or cystic fibrosis; chronic heart disease included chronic heart failure, ischaemic heart disease, and severe valve or congenital heart disease likely to require lifelong follow up. Chronic neurological conditions were separated into diseases with a likely cardiovascular aetiology (stroke, TIA, dementia) and conditions in which respiratory function may be compromised such as motor neurone disease, myasthenia gravis, multiple sclerosis, Parkinson’s disease, cerebral palsy, quadriplegia or hemiplegia, malignant primary brain tumour, and progressive cerebellar disease. Asplenia included splenectomy or a spleen dysfunction, including sickle cell disease. Other immunosuppressive conditions included HIV or a condition inducing permanent immunodeficiency ever diagnosed, or aplastic anaemia or temporary immunodeficiency recorded within the last year. Haematological malignancies were considered separately from other cancers to reflect the immunosuppression associated with haematological malignancies and their treatment. Kidney function was ascertained from the most recent serum creatinine measurement, where available, converted into estimated glomerular filtration rate (eGFR) using the Chronic Kidney Disease Epidemiology Collaboration (CKD-EPI) equation,^22^ with impaired kidney function defined as eGFR <60 mL/min/1.73m^2^. Raised blood pressure (BP) was defined as either a prior coded diagnosis of hypertension or the most recent recording indicating systolic BP ≥140 mmHg or diastolic BP ≥90 mmHg.

Asthma was grouped by use of oral corticosteroids as an indication of severity. Diabetes was grouped according to the most recent Hba1c measurement, where a measurement was available within the last 15 months, into controlled (Hba1c < 58 mmols/mol) and uncontrolled (Hba1c >= 58 mmols/mol). Cancer was grouped by time since the first diagnosis (within the last year, 2-<5 years, >5 years).

Other covariates considered as potential upstream risk factors were deprivation and ethnicity. Deprivation was measured by the Index of Multiple Deprivation (IMD, in quintiles, with higher values indicating greater deprivation), derived from the patient’s postcode at lower super output area level for a high degree of precision. Ethnicity was grouped into White, Black, Asian or Asian British, Mixed, or Other. The Sustainability and Transformation Partnership (STP, an NHS administrative region) of the patient’s general practice was included as an additional adjustment for geographical variation in infection rates across the country.

Information on all covariates were obtained from primary care records by searching TPP SystmOne records for specific coded data. TPP SystmOne allows users to work with the SNOMED-CT clinical terminology, using a GP subset of SNOMED-CT codes. This subset maps on to the native Read version 3 (CTV3) clinical coding system that SystmOne is built on. Medicines are entered or prescribed in a format compliant with the NHS Dictionary of Medicines and Devices (dm+d),^23^ a local UK extension library of SNOMED. Code lists for particular underlying conditions and medicines were compiled from a variety of sources. These include BNF codes from OpenPrescribing.net, published codelists for asthma,^24-26^ immunosuppression,^27-29^ psoriasis,^30^ SLE,^31^ RA^32,33^ and cancer,^34,35^ and Read Code 2 lists designed specifically to describe groups at increased risk of influenza infection.^36^ Read Code 2 lists were added to with SNOMED codes and cross-checked against NHS QOF registers, then translated into CTV3 with manual curation. Decisions on every code list were documented and final lists reviewed by at least two authors. Detailed information on compilation and sources for every individual codelist is available at https://codelists.opensafely.org/ and the lists are available for inspection and re-use by the broader research community.^37^

### Statistical Analysis

Patient numbers are depicted in figure 1. The Kaplan-Meier failure function by age group and sex are shown in figure 2. For each potential risk factor, a Cox proportional hazards model was fitted, with days in study as the timescale, stratified by geographic area (STP), and adjusted for sex and age modelled using restricted cubic splines. Violations of the proportional hazards assumption were explored by testing for a zero slope in the scaled Schoenfeld residuals. All potential risk factors, including age (again modelled as a spline), sex, BMI, smoking, index of multiple deprivation quintile, and comorbidities listed above were then included in a single multivariable Cox proportional hazards model, stratified by STP. Hazard ratios from the age/sex adjusted and fully adjusted models are reported with 95% confidence intervals. Models were also refitted with age group fitted as a categorical variable in order to obtain hazard ratios by age group.

**Figure 1.**
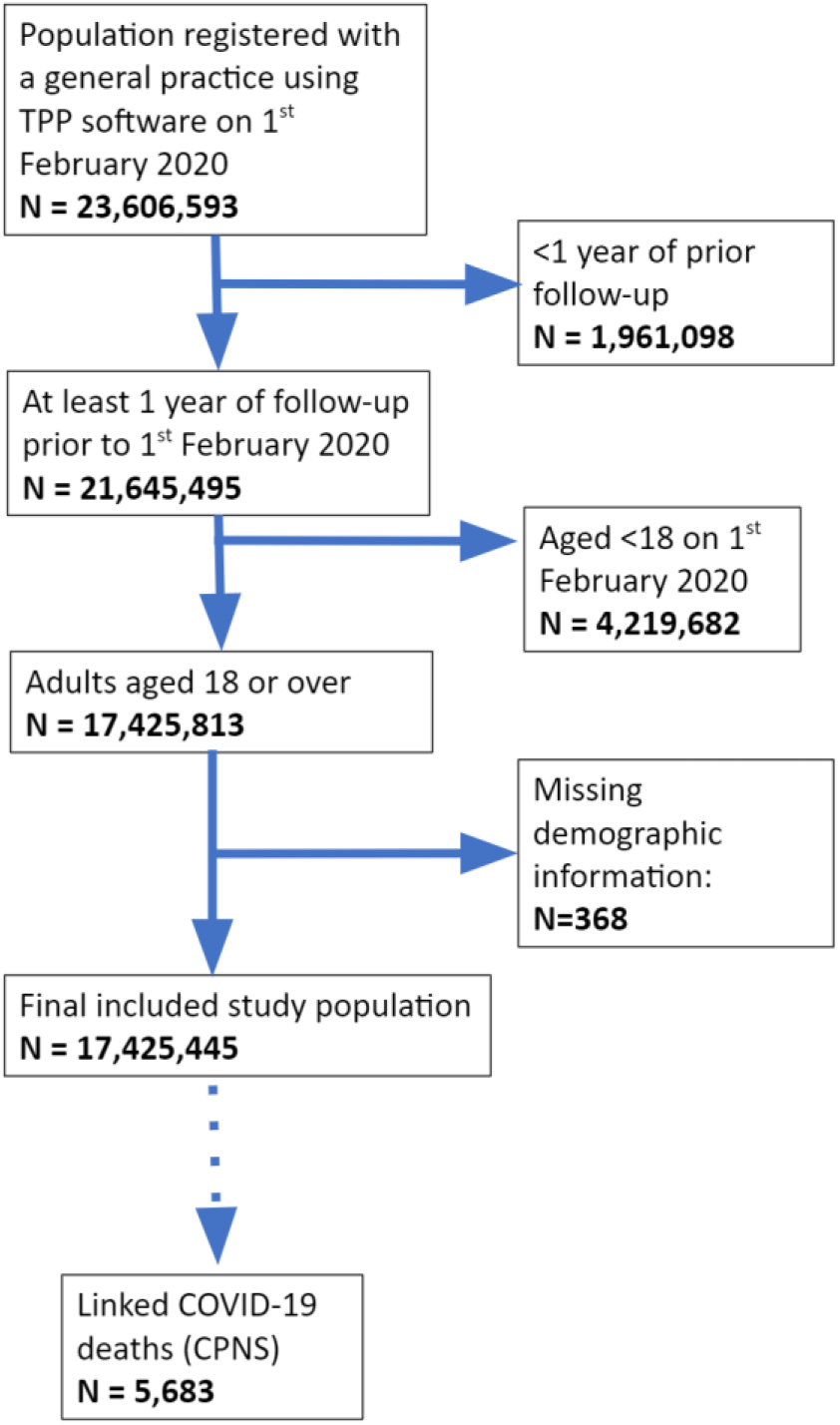
Flow diagram of cohort with numbers excluded at different stages and identification of cases for the main endpoints.

**Figure 2.**
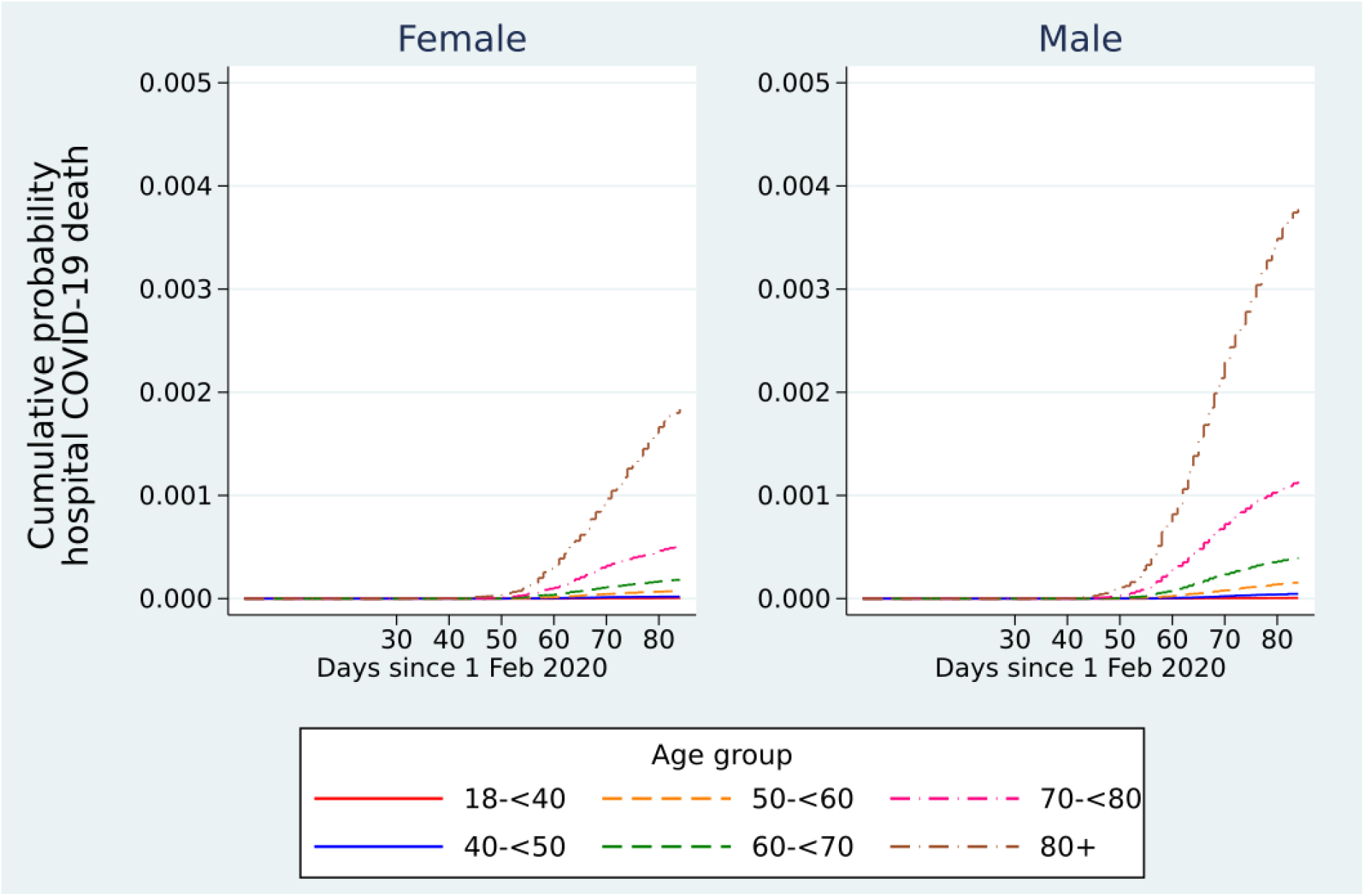
Kaplan-Meier plots for in-hospital COVID-19 death over time by age and sex.

In the primary analysis, those with missing BMI were assumed non-obese and those with missing smoking information were assumed to be nonsmokers on the assumption that both obesity and smoking would be likely to be recorded if present. A sensitivity analysis was run among those with complete BMI and smoking data only. Ethnicity was omitted from the main multivariable model due to 26% of individuals having missing data; hazard ratios for ethnicity were therefore obtained from a separate model among individuals with complete ethnicity only. Hazard ratios for other risk factors, adjusted for ethnicity, were also obtained from this model and are presented in the sensitivity analyses to allow assessment of the potential for confounding by ethnicity in the primary model. All multivariable models excluded the very small number of patients (<1%) with missing IMD.

The C-statistic was calculated as a measure of model discrimination. Due to computational time, this was estimated by randomly sampling 5000 patients without the outcome and calculating the C-statistic using the random sample and all patients who experienced the outcome, repeating this 10 times and taking the average C-statistic.

### Information governance and ethics

NHS England is the data controller; TPP is the data processor; and the key researchers on OpenSAFELY are acting on behalf of NHS England. This implementation of OpenSAFELY is hosted within the TPP environment which is accredited to the ISO 27001 information security standard and is NHS IG Toolkit compliant;^38,39^ patient data has been pseudonymised for analysis and linkage using industry standard cryptographic hashing techniques; all pseudonymised datasets transmitted for linkage onto OpenSAFELY are encrypted; access to the platform is via a virtual private network (VPN) connection, restricted to a small group of researchers, their specific machine and IP address; the researchers hold contracts with NHS England and only access the platform to initiate database queries and statistical models; all database activity is logged; only aggregate statistical outputs leave the platform environment following best practice for anonymisation of results such as statistical disclosure control for low cell counts.^40^ The OpenSAFELY research platform adheres to the data protection principles of the UK Data Protection Act 2018 and the EU General Data Protection Regulation (GDPR) 2016. In March 2020, the Secretary of State for Health and Social Care used powers under the UK Health Service (Control of Patient Information) Regulations 2002 (COPI) to require organisations to process confidential patient information for the purposes of protecting public health, providing healthcare services to the public and monitoring and managing the COVID-19 outbreak and incidents of exposure.^41^ Taken together, these provide the legal bases to link patient datasets on the OpenSAFELY platform. This study was approved by the Health Research Authority (REC reference 20/LO/0651) and by the LSHTM Ethics Board (reference 21863).

### Software and Reproducibility

Data management was performed using Python 3.8 and SQL, with analysis carried out using Stata 16.1 / Python. All code for data management and analysis is archived online at https://github.com/ebmdatalab/opensafely-risk-factors-research. All clinical and medicines codelists are openly available for inspection and reuse at https://codelists.opensafely.org/.

### Patient and Public Involvement

Patients were not formally involved in developing this specific study design. We have developed a publicly available website https://opensafely.org/ allowing any patient or member of the public to contact us regarding this study or the broader OpenSAFELY project. This feedback will be used to refine and prioritise our OpenSAFELY activities.

## Results

17,425,445 adults were included (Figure 1). Table 1 shows distributions of demographics and baseline comorbidities. 1,870,069 (11%) individuals had non-white ethnicities recorded. Missing data were present for body mass index (3,782,768, 22%), smoking status (725,323, 4%), ethnicity (4,592,377, 26%), IMD (142,166, 0.8%), and blood pressure (1,728,479, 10%). 5683 of the individuals had a COVID-19 hospital death recorded in CPNS.

**Table 1.** Cohort description with number of CPNS in-hospital deaths by potential risk factors.

|  | N (column %) | Number of CPNS Hospital deaths (% within stratum) |
| --- | --- | --- |
| **Total** | 17,425,445 (100.0) | 5683 (0.03) |
| **Age** |  |  |
| 18-<40 | 5,990,809 (34.4) | 40 (0.00) |
| 40-<50 | 2,875,561 (16.5) | 94 (0.00) |
| 50-<60 | 3,068,883 (17.6) | 355 (0.01) |
| 60-<70 | 2,405,327 (13.8) | 693 (0.03) |
| 70-<80 | 1,948,095 (11.2) | 1,560 (0.08) |
| 80+ | 1,136,770 (6.5) | 2,941 (0.26) |
| **Sex** |  |  |
| Female | 8,729,741 (50.1) | 2,098 (0.02) |
| Male | 8,695,704 (49.9) | 3,585 (0.04) |
| **BMI (kg/m<sup>2</sup>)** |  |  |
| <18.5 | 312,894 (1.8) | 161 (0.05) |
| 18.5-24.9 | 4,806,089 (27.6) | 1,467 (0.03) |
| 25-29.9 | 4,723,031 (27.1) | 1,663 (0.04) |
| 30-34.9 | 2,404,098 (13.8) | 1,164 (0.05) |
| 35-39.9 | 929,803 (5.3) | 467 (0.05) |
| ≥40 | 466,762 (2.7) | 257 (0.06) |
| Missing | 3,782,768 (21.7) | 504 (0.01) |
| **Smoking** |  |  |
| Never | 8,000,204 (45.9) | 1,734 (0.02) |
| Former | 5,737,545 (32.9) | 3,527 (0.06) |
| Current | 2,962,373 (17.0) | 393 (0.01) |
| *Missing* | 725,323 (4.2) | 29 (0.00) |
| **Ethnicity** |  |  |
| White | 10,962,999 (62.9) | 3,597 (0.03) |
| Mixed | 171,929 (1.0) | 39 (0.02) |
| Asian or Asian British | 1,030,890 (5.9) | 373 (0.04) |
| Black | 343,437 (2.0) | 158 (0.05) |
| Other | 323,813 (1.9) | 59 (0.02) |
| *Missing* | 4,592,377 (26.4) | 1,457 (0.03) |
| **IMD quintile** |  |  |
| 1 (least deprived) | 3,498,853 (20.1) | 948 (0.03) |
| 2 | 3,478,227 (20.0) | 1,040 (0.03) |
| 3 | 3,484,518 (20.0) | 1,101 (0.03) |
| 4 | 3,481,294 (20.0) | 1,246 (0.04) |
| 5 (most deprived) | 3,340,387 (19.2) | 1,316 (0.04) |
| *Missing* | 142,166 (0.8) | 32 (0.02) |
| **Blood pressure** |  |  |
| Normal | 3,845,356 (22.1) | 1,215 (0.03) |
| Elevated | 2,504,790 (14.4) | 971 (0.04) |
| High Stage 1 | 5,593,822 (32.1) | 1,796 (0.03) |
| High Stage 2 | 3,752,998 (21.5) | 1,688 (0.04) |
| *Missing* | 1,728,479 (9.9) | 13 (0.00) |
| High BP or diagnosed hypertension | 5,962,122 (34.2) | 4,204 (0.07) |
| **Comorbidities** |  |  |
| **Respiratory disease ex asthma** | 707,284 (4.1) | 1,274 (0.18) |
| **Asthma\*** |  |  |
| Present+recent ocs | 294,003 (1.7) | 201 (0.07) |
| Present, no recent ocs | 2,479,371 (14.2) | 710 (0.03) |
| **Chronic heart disease** | 1,173,443 (6.7) | 2,049 (0.17) |
| **Diabetes\*\*** |  |  |
| Uncontrolled<br>(HbA1c≥58 mmol/mol) | 489,297 (2.8) | 794 (0.16) |
| Controlled<br>(HbA1c<58 mmol/mol) | 1,043,176 (6.0) | 1,366 (0.13) |
| Present, no HbA1c | 195,243 (1.1) | 213 (0.11) |
| **Cancer (non-haematological)** |  |  |
| < 1 year ago | 80,334 (0.5) | 106 (0.13) |
| 1-4.9 years ago | 235,635 (1.4) | 247 (0.10) |
| ≥5 years ago | 545,223 (3.1) | 557 (0.10) |
| **Haematological malignancy** |  |  |
| < 1 year ago | 8,725 (0.1) | 27 (0.31) |
| 1-4.9 years ago | 27,925 (0.2) | 80 (0.29) |
| ≥5 years ago | 63,818 (0.4) | 103 (0.16) |
| **Liver disease** | 114,303 (0.7) | 111 (0.10) |
| **Stroke/dementia** | 373,968 (2.1) | 999 (0.27) |
| **Other neurology dis** | 171,975 (1.0) | 313 (0.18) |
| **Kidney disease** | 1,090,760 (6.3) | 2,541 (0.23) |
| **Organ transplant** | 20,130 (0.1) | 49 (0.24) |
| **Spleen diseases** | 28,160 (0.2) | 23 (0.08) |
| **Rheumatoid/Lupus/ Psoriasis** | 885,284 (5.1) | 533 (0.06) |
| **Other immunosuppressive condition** | 280,783 (1.6) | 36 (0.01) |

\* ocs = oral corticosteroid use, recent defined as <1 year before baseline, \*\* classification by HbA1c based on measures within 15 months before baseline.

The overall cumulative incidence of COVID-19 hospital death at 80 days from the study start date was <0.01% in those aged 18-39 years, rising to 0.35% and 0.17% in men and women respectively aged ≥80 years, with a trend by age (Figure 2).

Associations between patient-level factors and risk of COVID-19 hospital death are shown in Table 2 and Figure 3. Increasing age was strongly associated with risk, with the ≥80 years age group having more than 12-fold increased risk compared with those aged 50-59 years (fully adjusted HR 12.64; 95% CI 11.19-14.28). With age fitted as a flexible spline, an approximately log-linear relationship was observed (appendix Figure A1), equivalent to risk increasing exponentially with age. Male gender was associated with a doubling of risk (fully adjusted HR 1.99, 1.88-2.10).

**Table 2.** Hazard Ratios (HRs) and 95% confidence intervals (CI) for in-hospital COVID-19 death.

|  | CPNS Death HR (95% CI) |  |
| --- | --- | --- |
|  | Age-sex adj | Fully adj |
| **Age** |  |  |
| 18-<40 | 0.05 (0.04-0.08) | 0.07 (0.05-0.10) |
| 40-<50 | 0.27 (0.21-0.34) | 0.31 (0.25-0.39) |
| 50-<60 | 1.00 (ref) | 1.00 (ref) |
| 60-<70 | 2.61 (2.29-2.96) | 2.09 (1.84-2.38) |
| 70-<80 | 7.61 (6.78-8.54) | 4.77 (4.23-5.38) |
| 80+ | 26.27 (23.52-29.33) | 12.64 (11.19-14.28) |
| **Sex** |  |  |
| Female | 1.00 (ref) | 1.00 (ref) |
| Male | 2.24 (2.12-2.36) | 1.99 (1.88-2.10) |
| **BMI** |  |  |
| Not obese | 1.00 (ref) | 1.00 (ref) |
| Obese class I (30-34.9kg/m<sup>2</sup>) | 1.57 (1.47-1.68) | 1.27 (1.18-1.36) |
| Obese class II (35-39.9kg/m<sup>2</sup>) | 2.01 (1.82-2.21) | 1.56 (1.41-1.73) |
| Obese class III (≥40 kg/m<sup>2</sup>) | 2.97 (2.62-3.38) | 2.27 (1.99-2.58) |
| **Smoking** |  |  |
| Never | 1.00 (ref) | 1.00 (ref) |
| Ex-smoker | 1.80 (1.70-1.90) | 1.25 (1.18-1.33) |
| Current | 1.25 (1.12-1.40) | 0.88 (0.79-0.99) |
| **Ethnicity\*** |  |  |
| White | 1.00 (ref) | 1.00 (ref) |
| Mixed | 1.83 (1.33-2.51) | 1.64 (1.19-2.26) |
| Asian or Asian British | 1.95 (1.73-2.18) | 1.62 (1.43-1.82) |
| Black | 2.17 (1.84-2.57) | 1.71 (1.44-2.02) |
| Other | 1.34 (1.03-1.74) | 1.33 (1.03-1.73) |
| **IMD quintile** |  |  |
| 1 (least deprived) | 1.00 (ref) | 1.00 (ref) |
| 2 | 1.18 (1.08-1.29) | 1.13 (1.04-1.24) |
| 3 | 1.35 (1.23-1.47) | 1.23 (1.13-1.35) |
| 4 | 1.70 (1.56-1.86) | 1.49 (1.37-1.63) |
| 5 (most deprived) | 2.13 (1.95-2.33) | 1.75 (1.60-1.91) |
| **Blood pressure** |  |  |
| Normal | 1.00 (ref) | 1.00 (ref) |
| High, or diagnosed hyper-tension | 1.22 (1.15-1.30) | 0.95 (0.89-1.01) |
| **Co-morbidities** |  |  |
| **Respiratory disease ex asthma** | 2.35 (2.21-2.50) | 1.78 (1.67-1.90) |
| **Asthma (vs none)\*<sup>2</sup>** |  |  |
| With no recent OCS use | 1.23 (1.14-1.33) | 1.11 (1.02-1.20) |
| With recent OCS use | 1.70 (1.48-1.96) | 1.25 (1.08-1.44) |
| **Chronic heart disease** | 2.01 (1.90-2.13) | 1.27 (1.20-1.35) |
| **Diabetes (vs none)\*<sup>3</sup>** |  |  |
| Controlled (HbA1c<58 mmol/mol) | 2.02 (1.89-2.16) | 1.50 (1.40-1.60) |
| Uncontrolled (HbA1c≥58 mmol/mol) | 3.61 (3.34-3.90) | 2.36 (2.18-2.56) |
| No recent HbA1c measure | 2.35 (2.04-2.70) | 1.87 (1.63-2.16) |
| **Cancer (non-haematological, vs none)** |  |  |
| Diagnosed < 1 year ago | 1.83 (1.51-2.21) | 1.56 (1.29-1.89) |
| Diagnosed 1-4.9 years ago | 1.39 (1.22-1.58) | 1.19 (1.04-1.35) |
| Diagnosed ≥5 years ago | 1.03 (0.94-1.12) | 0.97 (0.88-1.06) |
| **Haematological malignancy (vs none)** |  |  |
| Diagnosed < 1 year ago | 4.03 (2.76-5.88) | 3.52 (2.41-5.14) |
| Diagnosed 1-4.9 years ago | 3.59 (2.88-4.48) | 3.12 (2.50-3.89) |
| Diagnosed ≥5 years ago | 2.13 (1.76-2.59) | 1.88 (1.55-2.29) |
| **Liver disease** | 2.34 (1.94-2.83) | 1.61 (1.33-1.95) |
| **Stroke/dementia** | 2.34 (2.18-2.51) | 1.79 (1.67-1.93) |
| **Other neurological** | 2.94 (2.62-3.30) | 2.46 (2.19-2.76) |
| **Kidney disease** | 2.19 (2.06-2.32) | 1.72 (1.62-1.83) |
| **Organ transplant** | 7.79 (5.88-10.33) | 4.27 (3.20-5.70) |
| **Spleen diseases** | 1.82 (1.21-2.74) | 1.41 (0.93-2.12) |
| **Rheumatoid/ Lupus/ Psoriasis** | 1.35 (1.24-1.48) | 1.23 (1.12-1.35) |
| **Other immunosuppressive condition** | 2.02 (1.45-2.81) | 1.69 (1.21-2.34) |

Models adjusted for age using a 4-knot cubic spline age spline, except for estimation of age group effects.
\*Ethnicity effect estimated from a model restricted to those with recorded ethnicity. OCS = oral corticosteroids.
\*<sup>2</sup>Recent OCS use defined as in the year before baseline. \*<sup>3</sup>HbA1c classification based on latest measure within 15 months before baseline.

**Figure 3.**
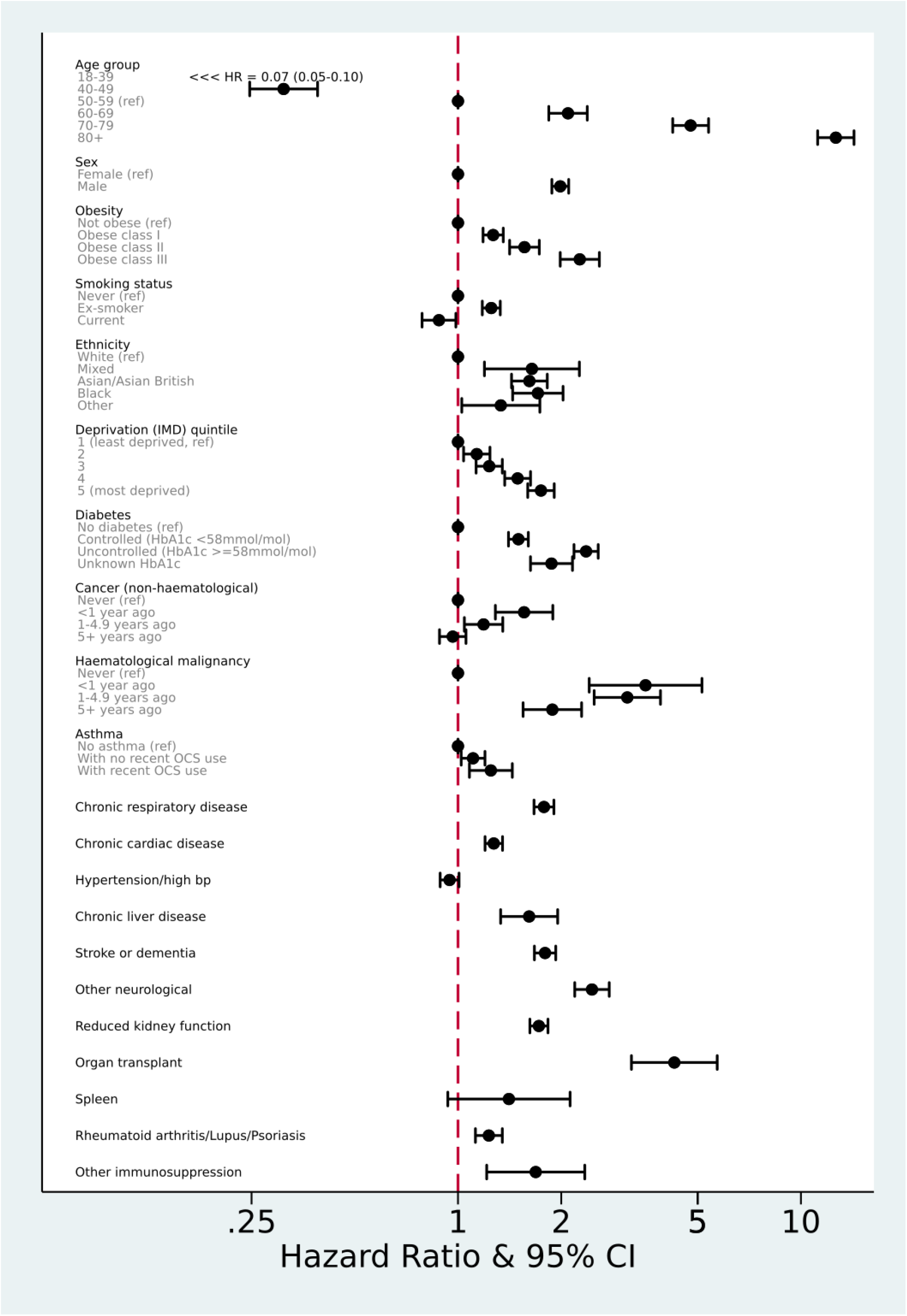
Estimated Hazard Ratios (shown on a log scale) for each potential risk factor from a multivariable Cox model. Obese class I:30-34.9kg/m2, class II: 35-39.9kg/m2, class III: >=40kg/m2. OCS = oral corticosteroid. All HRs are adjusted for all other factors listed other than ethnicity. Ethnicity estimates are from a separate model among those with complete ethnicity data, and are fully adjusted for other covariates.

All non-white ethnic groups had higher risk than those with white ethnicity: HRs adjusted for age and sex only ranged from 1.83-2.17 for Black, Asian/Asian British and mixed ethnicities compared to white; these attenuated to 1.62-1.71 on adjustment for all included risk factors. Increasing risks were seen with increasing levels of deprivation, and with increasing levels of obesity (BMI >40 fully adjusted HR 2.27, 95% CI 1.99-2.58).

Both current and former smoking were associated with higher risk in models adjusted for age and sex only, but in the fully adjusted model there was weak evidence of a slightly lower risk in current smokers (fully adjusted HRs 0.88, CI 0.79-0.99). In post-hoc analyses we added individual covariates to the model with age, sex and smoking to explore this further: the change in HR appeared to be largely driven by adjustment for chronic respiratory disease (HR 0.93, 0.83-1.04 after adjustment) and deprivation (HR 0.98, 0.88-1.10 after adjustment). Other individual adjustments did not remove the positive association between current smoking and outcome. We also explored confounding by ethnicity, which was not adjusted in the primary model: among those with complete ethnicity, the current smoking HR adjusted for all variables except ethnicity was similar to in the full study population (0.88, 0.78-1.01) but this moved towards the null on adjustment for ethnicity (HR 0.94, 0.82-1.07).

Most comorbidities were associated with higher risk of COVID-19 hospital death, including diabetes (with a greater HR for those with recent HbA1c >= 58 mmol/mol), asthma (with a greater HR for those with recent use of an oral corticosteroid), respiratory disease, chronic heart disease, liver disease, stroke/dementia, other neurological diseases, reduced kidney function, autoimmune diseases (rheumatoid arthritis, lupus or psoriasis) and other immunosuppressive conditions, as per Table 2. Those with a history of haematological malignancy were at >3-fold increased risk up to 5 years from diagnosis, and nearly double the risk thereafter. For other cancers, HRs were smaller and risk increases were largely observed among those diagnosed in the last year. There was no association between hypertension (defined as a recorded diagnosis, or high blood pressure at the last measurement) and outcome (HR 0.95, 0.89-1.01). However, in sensitivity analyses, diagnosed hypertension was associated with slightly increased risk (HR 1.07, 1.00-1.15) while high blood pressure (≥140/90 mmHg) at the most recent measurement was associated with lower risk (HR 0.61, 0.56-0.67).

The average C-statistic was 0.78. Sensitivity analyses are shown in Table A1 (appendix). Results were similar when restricted to those with complete BMI and smoking information, and when adjusted for ethnicity, among those with complete data. Violation of proportional hazards was detected in the primary model (p<0.001), so a further sensitivity analysis was run with earlier administrative censoring at 6th April 2020, since social distancing measures introduced across the UK in late March 2020 would not have been expected to impact on mortality rates at that time. There was no evidence of non-proportional hazards in this analysis (p=0.56). The overall pattern of results was similar to the primary model, though most HRs were somewhat larger in magnitude in the analysis restricted to this earlier period, while the effect of increasing deprivation appeared to be smaller (appendix Table A1).

## Discussion

### Summary

We have successfully delivered a secure analytics platform operating across almost 24 million patient records for the Covid-19 emergency, and used this to identify, quantify, and further explore a range of risk factors for death in hospital from COVID-19 in the largest cohort study conducted by any country to date. Most comorbidities we studied were associated with increased risk, including cardiovascular disease, diabetes, respiratory disease including asthma, obesity, history of haematological malignancy or recent other cancer, kidney, liver, neurological and autoimmune conditions. People from Asian and black groups had a substantially higher risk of death from COVID-19, only partially attributable to co-morbidity, deprivation or other risk factors. Deprivation is also a major risk factor, which was only partly attributable to co-morbidity or other risk factors.

### Strengths and weaknesses

The greatest strengths of this study were speed and size. By building a secure analytics platform across routinely collected live clinical data stored in situ we have been able to produce timely results from the current records of approximately 40% of the English population in response to a global health emergency. This scale allowed us to work with more precision, on rarer exposures, on multiple risk factors, and to detect important signals as early as possible in the course of the pandemic. The scale of our platform will shortly expand further, and we will report updated analyses over time. Another key strength is our use of open methods: we pre-specified our analysis plan and have shared our full analytic code and all code lists for review and re-use. We ascertained patients’ demographics, medications and comorbidities from their full pseudonymised longitudinal primary care records, providing substantially more detailed information than is available in hospital records or data recorded at time of admission alone, and on the total population at risk rather than the selected subset presenting for treatment in hospital. Linkage to ONS allowed censoring of data in the control population for patients who had died outside hospital or from other causes. Analyses were stratified by area to account for known geographical differences in incidence of COVID-19.

We also identify important limitations. Using CPNS data alone relies on hospitals completing a new return under emergency conditions; furthermore COVID-19 deaths among people with false-negative tests and those untested may not have been included; we will validate CPNS against ONS data (which has a longer delay to reporting) as more cases arise. This initial analysis was focused on in-hospital death: our findings therefore reflect both an individual’s risk of infection, and their risk of dying once infected. We will explore patient trajectories in future research using test results and A&E presentation data which are now being linked on the OpenSAFELY platform.

Censoring patients at date of death from other causes, or outside hospital, was only possible until 16th April 2020, 9 days prior to study end. Rather than ending the study earlier, greatly reducing case numbers, we end at 25 April, acknowledging that censoring would be incomplete for the last 9 days. Consequently a small number of the sickest individuals, who died after being discharged from hospital, will have remained in the “at risk” group over the last few days of study time when they should be censored. This would most likely apply to those with risk factors present, and therefore attenuate HRs, but any impact is likely to be small: a sensitivity analysis with all data censored at 6 April showed minimal differences.

Our analysis to date covers 40% of the population, but may not yet be fully representative as it currently includes only practices using the EHR software SystmOne: there is substantial geographic variation in choice of EHR system and in London, where many earlier COVID-19 cases occurred, only 17% of general practices use SystmOne. Additionally it has been shown that the user interface of electronic health records can affect prescribing of certain medicines^42-44^ so it is possible that coding of conditions may vary between systems; again we will evaluate this further with more data.

Primary care records, though detailed and longitudinal, can be incomplete for data on risk factors and other covariates. In particular, ethnicity was not recorded in approximately 26% of patients included; prior research has shown that when ethnicity is recorded in EHR its distribution is very similar to that in census ethnicity data.^45^ Obesity and smoking were assumed absent if missing; patients with missing creatinine and blood pressure measurements were included in the categories denoting no evidence of reduced kidney function or high blood pressure respectively. We undertook a number of sensitivity analyses to assess robustness of these approaches, hazard ratios were similar across analyses. Deprivation score may be inaccurate for any patients without an up-to-date address, but this is unlikely to introduce a strong bias in any particular direction.

Deviations from proportional hazards were detected. This could be partly or wholly due to the very large numbers included meaning small deviations are statistically significant, or due to unmeasured covariates. However, it may have been due to rapid changes in social behaviours following government advice on social distancing, shielding, and changes in the pattern and burden of infection across the UK, which may also have affected different patient groups differentially. A sensitivity analysis with early censoring at 6th April 2020 (before social distancing and shielding measures would be likely to affect mortality) showed no evidence of non-proportional hazards (p=0.56) and similar results to the primary model, but with larger hazard ratios for several risk factors. This is consistent with the hypothesis that the most at-risk patients may have been more compliant with social distancing and shielding policies introduced later. In contrast, the effect of increased deprivation appeared to be smaller in the earlier period, suggesting that the risk associated with deprivation may have increased over time. Subsequent analyses will explore different analytical approaches, including fitting interactions with time, and using accelerated failure time models, to further explore changes before and after national initiatives around COVID-19.

### Findings in Context

Our findings on age and gender are consistent with patterns observed worldwide in smaller studies on patients infected and/or dying from COVID-19. Compared to white people, people of Asian and Black ethnic origin were found to be at a higher risk of death. Non-white ethnicity has previously been found to be associated with increased COVID-19 infection and poor outcomes.^12,13,46^ Commentators and researchers have reasonably speculated that this might be due to higher prevalence of medical problems such as cardiovascular disease or diabetes among BME people, or higher deprivation. Our findings, based on more detailed data, show that this is only a small part of the excess risk. Other possible explanations for increased risk among BME groups relate to higher infection risk, including over-representation in ‘front-line” professions with higher exposure to infection, or higher household density. Addressing these questions will likely entail bespoke data collection on, for example, occupation among cases and controls.

We also found a consistent pattern of increasing risk with greater deprivation, with the most deprived quintile having a HR of 1.75 compared to the least deprived, consistent with recent national statistics.^47^ Again, contrary to prior speculation, very little of the increased risk associated with deprivation was explained by pre-existing disease or clinical risk factors, suggesting that other social factors increase the risk of COVID-19 infection or death from infection.

We found increased risk for the major included co-morbidities. The ISARIC study describing 16,749 hospitalised UK patients with COVID-19 also indicated increased risk of death among hospitalised patients with cardiac, pulmonary and kidney disease, malignancy and dementia.^6^ Cardiovascular disease, hypertension, diabetes, respiratory disease, and cancers were all associated with increased risk of death in a large Chinese study describing 44,672 confirmed cases, but which lacked age-correction.^5^

Of particular note in our results is the association of asthma with higher risk of COVID-19 hospital death, with the HR increasing further for those having a recent oral corticosteroid (indicating greater severity of disease). This contrasts with previous findings: in several countries, asthma and other chronic respiratory diseases are underrepresented in hospitalised patients;^48^ and among the first few hundred cases in UK, a protective association with asthma was observed, although presence of asthma was ascertained differently for cases and controls which could be a source of bias.^36^ The ISARIC study reported 14% of hospitalised patients having asthma but no increased risk of death.^6^ However, in both the UK and China, COPD did appear to confer increased risk of death among hospitalised or confirmed cases, respectively.^5,6^ Our study design combines both risk of infection *and* risk of death once infected; it is also possible that our methodology captured more people with asthma and was better able to delineate more severe asthma than previous studies based on hospital records. We found no association between hypertension and death where hypertension was defined as a recorded diagnosis or high blood pressure at last measurement (HR 0.96, 0.9-1.02). However when separated out in sensitivity analyses diagnosed hypertension was associated with slightly increased risk, whilst a high blood pressure measurement was associated with slightly reduced risk. Hypertension is very strongly associated with age and although we adjusted for this, disentangling the effects of each is difficult.

We showed increasing risk of death with degree of obesity: fully-adjusted HR 1.27 for BMI 30-34.9 kg/m^2^, increasing to 2.27 for BMI >=40 kg/m^2^. Previous studies have shown increased risk with obesity among hospitalised patients: ISARIC, based on hospital survey data (2,212 deaths), found a HR of 1.37 for death associated with clinician-reported obesity among 16,749 hospitalised patients, after age and sex correction (95% CI 1.16-1.63)^6^; obesity was also reported as a risk factor for treatment escalation in smaller studies in France and New York.^7,8^

We found some evidence of increased risks in former smokers. In current smokers there was a slight protective effect, which was removed when fully adjusted for ethnicity. The risks associated with smoking have been disputed, with increased risks initially reported, but some more recent reports finding that smokers are under-represented in those with more severe disease, and a potential protective mechanism for nicotine has been suggested^9,49^: smoking prevalence was lower than expected among hospitalised patients in China,^10^ France^11^ and the USA.^50^ Even if smoking does have a small protective effects against COVID-19, this would still be massively outweighed by the well-established adverse health effects of smoking.

### Policy Implications and Interpretation

The UK has a policy of recommending shielding (i.e. minimising face to face contact) for groups identified as being extremely vulnerable to COVID-19 on the basis of pre-existing medical conditions. We were able to evaluate the association between most of these conditions and death from COVID-19, and confirm that people with these conditions do have substantially increased mortality risk, supporting the shielding strategy. We have demonstrated - for the first time - that only a small part of the substantially increased risks of death from COVID-19 among nonwhite groups and among people living in more deprived areas can be attributed to existing disease. Improved strategies to protect people in these groups from COVID-19 need urgent consideration.

The UK has an unusually large volume of very detailed longitudinal patient data, especially through primary care. We believe the UK has a responsibility to the global community to make good use of this data, securely, and to the highest scientific standards. OpenSAFELY demonstrates the value of this data in practice. We will enhance the OpenSAFELY platform to further inform the global response to the COVID-19 emergency.

### Future Research

The underlying causes of higher risk among those from non-white backgrounds, and deprived areas, require further exploration; we would suggest collecting data on occupational exposure and living conditions as first steps. The statistical power offered by our approach means that associations with less common risk factors can be robustly assessed in more detail, at the earliest possible date, as the pandemic progresses. We will therefore update our findings and address smaller risk groups as new cases arise over time. The open source re-usable codebase on OpenSAFELY supports rapid, secure and collaborative development of new analyses: we are currently conducting expedited studies on the impact of various medical treatments and population interventions on the risk of COVID-19 infection, ITU admission, and death, alongside other observational analyses. OpenSAFELY is rapidly scalable for additional NHS patients’ records, with new data sources progressing.

### Conclusion

We report early data on risk factors for death from COVID-19 using an unprecedented scale of 17 million patients’ detailed primary care records in the context of a global health emergency; we will update our findings as new data arises.

## Data Availability

All code for data management and analysis is archived online at https://github.com/ebmdatalab/opensafely-risk-factors-research. All clinical and medicines codelists are openly available for inspection and reuse at https://codelists.opensafely.org/.

https://codelists.opensafely.org/

## Acknowledgements

We are very grateful for all the support received from the TPP Technical Operations team throughout this work; for generous assistance from the information governance and database teams at NHS England / NHSX; and for additional discussions on disease characterisation, codelists, and methodology with Henry Drysdale, Brian Nicholson, Nick DeVito, Will Hulme, Jess Morley, and Jenni Quint.

## Ethical approval

This study was approved by the Health Research Authority (REC reference 20/LO/0651) and by the LSHTM Ethics Board (ref 21863). No further ethical or research governance approval was required by the University of Oxford but copies of the approval documents were reviewed and held on record.

## Guarantor

BG/LS.

## Contributorship

BG conceived the platform and the approach; BG and LS led the project overall and are guarantors; SB led the software; EW KB led the statistical analysis; CM AW led on codelists and implementation; AM led on IG; Contributions are as follows: Data curation CB JP JC SH SB DE PI CM; Analysis EW KB AW CM; Funding acquisition BG LS; information governance AM BG CB JP; Methodology EW KB AW BG LS CB JP JC SH SB DE PI CM RP; Disease category conceptualisation and codelists CM AW PI SB DE CB JC JP SH HD HC KB SB AM BM LT ID HM RM HF JQ; Ethics approval HC EW LS BG; Project administration CM HC CB SB AM LS BG; Resources BG LS FH; Software SB DE PI AW CM CB FH JC SH; Supervision BG LS SB; Writing (original draft) HC EW KB BM CM AM BG LS; Writing (review & editing) CB CM HC EW KB SB AM BM LT ID HM RM AW SE. All authors were involved in design and conceptual development and reviewed and approved the final manuscript.

## Appendix

**Figure A1.**
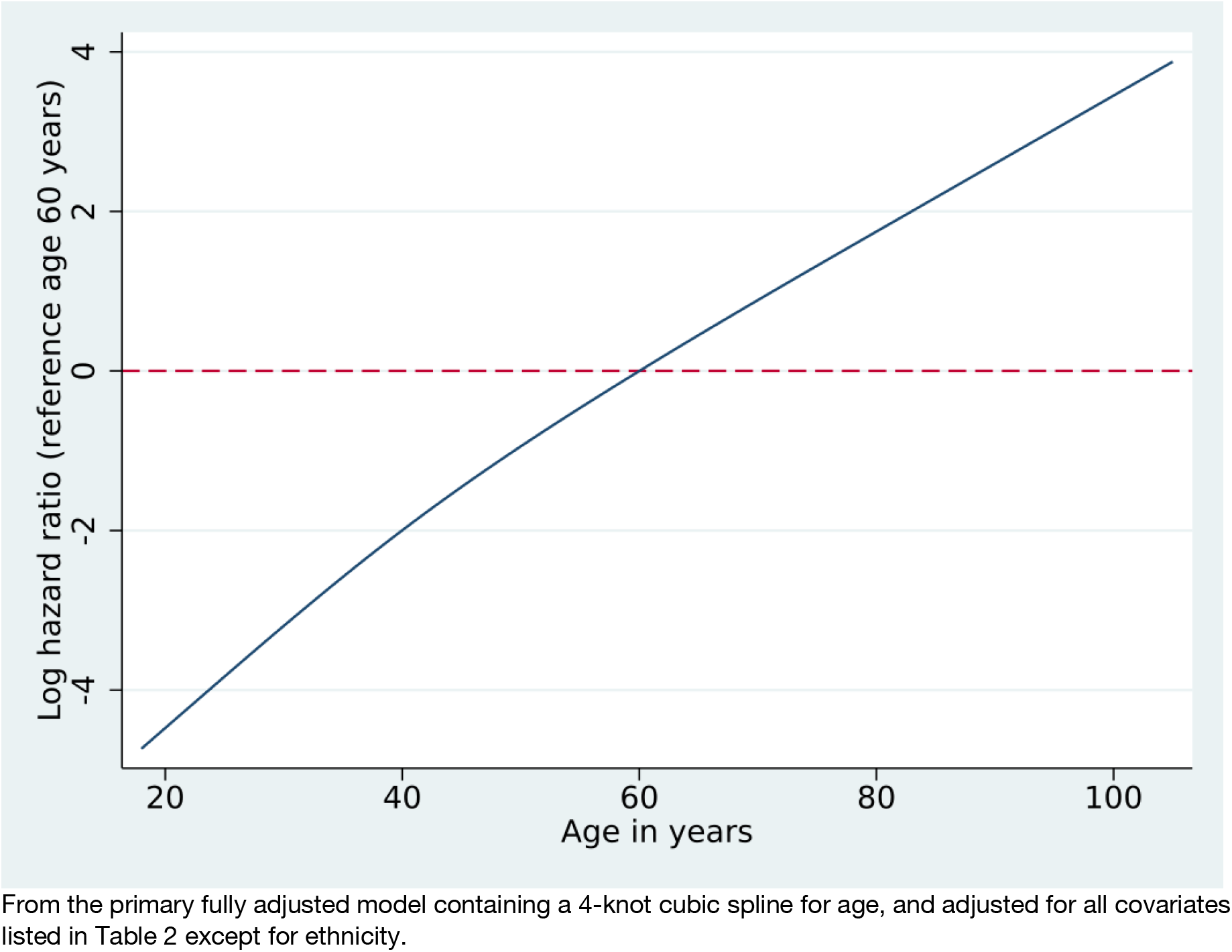
Estimated log hazard ratio by age in years

**Table A1.** Hazard Ratios (HRs) and 95% confidence intervals (CI) in sensitivity analyses

|  | Fully adjusted HR and 95% CI |  |  |  |
| --- | --- | --- | --- | --- |
|  | Primary analysis | Early censoring at 6/4/2020 | Restricted to those with complete BMI /smoking | Adjusted for ethnicity in those where recorded |
| **Age** |  |  |  |  |
| 18-<40 | 0.07 (0.05-0.10) | 0.08 (0.05-0.13) | 0.09 (0.06-0.13) | 0.08 (0.06-0.11) |
| 40-<50 | 0.31 (0.25-0.39) | 0.31 (0.21-0.46) | 0.32 (0.25-0.41) | 0.30 (0.23-0.38) |
| 50-<60 | 1.00 (ref) | 1.00 (ref) | 1.00 (ref) | 1.00 (ref) |
| 60-<70 | 2.09 (1.84-2.38) | 2.35 (1.90-2.91) | 2.12 (1.85-2.44) | 2.07 (1.79-2.39) |
| 70-<80 | 4.77 (4.23-5.38) | 5.55 (4.54-6.77) | 4.84 (4.25-5.50) | 4.81 (4.20-5.51) |
| 80+ | 12.64 (11.19-14.28) | 13.43 (10.95-16.45) | 12.76 (11.18-14.55) | 12.04 (10.47-13.84) |
| **Sex** |  |  |  |  |
| Female | 1.00 (ref) | 1.00 (ref) | 1.00 (ref) | 1.00 (ref) |
| Male | 1.99 (1.88-2.10) | 2.18 (1.99-2.38) | 2.06 (1.94-2.19) | 1.93 (1.80-2.06) |
| **BMI** |  |  |  |  |
| Not obese | 1.00 (ref) | 1.00 (ref) | 1.00 (ref) | 1.00 (ref) |
| Obese class I (30-34.9kg/m<sup>2</sup>) | 1.27 (1.18-1.36) | 1.39 (1.25-1.54) | 1.32 (1.23-1.41) | 1.28 (1.18-1.38) |
| Obese class II (35-39.9kg/m<sup>2</sup>) | 1.56 (1.41-1.73) | 1.62 (1.39-1.90) | 1.64 (1.48-1.81) | 1.60 (1.43-1.80) |
| Obese class III (≥40 kg/m<sup>2</sup>) | 2.27 (1.99-2.58) | 2.45 (2.00-3.01) | 2.40 (2.10-2.74) | 2.28 (1.96-2.65) |
| **Smoking** |  |  |  |  |
| Never | 1.00 (ref) | 1.00 (ref) | 1.00 (ref) | 1.00 (ref) |
| Ex-smoker | 1.25 (1.18-1.33) | 1.33 (1.21-1.47) | 1.24 (1.17-1.33) | 1.32 (1.23-1.42) |
| Current | 0.88 (0.79-0.99) | 0.84 (0.70-1.01) | 0.92 (0.81-1.03) | 0.94 (0.82-1.07) |
| **Ethnicity\*** |  |  |  |  |
| **White** | 1.00 (ref) | 1.00 (ref) | 1.00 (ref) | 1.00 (ref) |
| Mixed | 1.64 (1.19-2.26) | 1.13 (0.62-2.05) | 1.58 (1.12-2.22) | 1.64 (1.19-2.26) |
| Asian or Asian British | 1.62 (1.43-1.82) | 1.77 (1.48-2.13) | 1.69 (1.49-1.91) | 1.62 (1.43-1.82) |
| Black | 1.71 (1.44-2.02) | 1.90 (1.48-2.45) | 1.69 (1.42-2.02) | 1.71 (1.44-2.02) |
| Other | 1.33 (1.03-1.73) | 1.81 (1.28-2.57) | 1.41 (1.07-1.84) | 1.33 (1.03-1.73) |
| **IMD quintile** |  |  |  |  |
| **1 (least deprived)** | 1.00 (ref) | 1.00 (ref) | 1.00 (ref) | 1.00 (ref) |
| 2 | 1.13 (1.04-1.24) | 1.01 (0.88-1.16) | 1.12 (1.02-1.23) | 1.19 (1.07-1.33) |
| 3 | 1.23 (1.13-1.35) | 1.04 (0.91-1.20) | 1.23 (1.12-1.35) | 1.26 (1.13-1.40) |
| 4 | 1.49 (1.37-1.63) | 1.27 (1.11-1.46) | 1.48 (1.35-1.62) | 1.53 (1.38-1.70) |
| **5 (most deprived)** | 1.75 (1.60-1.91) | 1.49 (1.29-1.71) | 1.72 (1.57-1.89) | 1.70 (1.53-1.89) |
| **Blood pressure** |  |  |  |  |
| **Normal** | 1.00 (ref) | 1.00 (ref) | 1.00 (ref) | 1.00 (ref) |
| High, or diagnosed hypertension | 0.95 (0.89-1.01) | 0.94 (0.85-1.05) | 0.94 (0.88-1.01) | 0.97 (0.90-1.05) |
| **Co-morbidities** |  |  |  |  |
| **Respiratory disease ex asthma** | 1.78 (1.67-1.90) | 1.97 (1.77-2.18) | 1.74 (1.62-1.86) | 1.79 (1.66-1.93) |
| **Asthma (vs none)\*<sup>2</sup>** |  |  |  |  |
| With no recent OCS use | 1.11 (1.02-1.20) | 1.14 (1.01-1.29) | 1.10 (1.02-1.20) | 1.03 (0.94-1.13) |
| With recent OCS use | 1.25 (1.08-1.44) | 1.39 (1.12-1.73) | 1.22 (1.05-1.42) | 1.24 (1.06-1.46) |
| **Chronic heart disease** | 1.27 (1.20-1.35) | 1.33 (1.22-1.46) | 1.27 (1.19-1.35) | 1.27 (1.19-1.36) |
| **Diabetes (vs none)\*<sup>3</sup>** |  |  |  |  |
| Controlled (HbA1c<58 mmol/mol) | 1.50 (1.40-1.60) | 1.48 (1.33-1.65) | 1.47 (1.37-1.57) | 1.47 (1.36-1.59) |
| Uncontrolled (HbA1c≥58 mmol/mol) | 2.36 (2.18-2.56) | 2.57 (2.27-2.91) | 2.30 (2.12-2.50) | 2.23 (2.03-2.45) |
| No recent HbA1c measure | 1.87 (1.63-2.16) | 1.68 (1.33-2.12) | 1.85 (1.60-2.15) | 1.91 (1.63-2.24) |
| **Cancer (non-haematological, vs none)** |  |  |  |  |
| Diagnosed < 1 year ago | 1.56 (1.29-1.89) | 1.51 (1.10-2.05) | 1.52 (1.24-1.86) | 1.68 (1.36-2.09) |
| Diagnosed 1-4.9 years ago | 1.19 (1.04-1.35) | 1.36 (1.13-1.65) | 1.20 (1.05-1.37) | 1.21 (1.04-1.40) |
| Diagnosed ≥5 years ago | 0.97 (0.88-1.06) | 0.92 (0.79-1.06) | 0.96 (0.87-1.05) | 1.02 (0.92-1.13) |
| **Haematological malignancy (vs none)** |  |  |  |  |
| Diagnosed < 1 year ago | 3.52 (2.41-5.14) | 2.60 (1.30-5.22) | 3.77 (2.58-5.50) | 3.30 (2.10-5.18) |
| Diagnosed 1-4.9 years ago | 3.12 (2.50-3.89) | 3.67 (2.66-5.06) | 3.03 (2.40-3.83) | 3.42 (2.67-4.38) |
| Diagnosed ≥5 years ago | 1.88 (1.55-2.29) | 1.64 (1.18-2.28) | 1.90 (1.55-2.33) | 1.84 (1.46-2.32) |
| **Liver disease** | 1.61 (1.33-1.95) | 1.86 (1.40-2.47) | 1.59 (1.30-1.93) | 1.61 (1.30-2.00) |
| **Stroke/dementia** | 1.79 (1.67-1.93) | 1.61 (1.43-1.81) | 1.78 (1.65-1.92) | 1.75 (1.61-1.90) |
| **Other neurological** | 2.46 (2.19-2.76) | 2.28 (1.88-2.76) | 2.38 (2.10-2.69) | 2.41 (2.11-2.76) |
| **Kidney disease** | 1.72 (1.62-1.83) | 1.75 (1.58-1.92) | 1.71 (1.60-1.82) | 1.76 (1.64-1.89) |
| **Organ transplant** | 4.27 (3.20-5.70) | 2.62 (1.51-4.57) | 4.44 (3.31-5.96) | 4.59 (3.36-6.27) |
| **Spleen diseases** | 1.41 (0.93-2.12) | 1.87 (1.06-3.30) | 1.25 (0.79-1.96) | 1.43 (0.90-2.27) |
| **Rheumatoid/Lupus/Psoriasis** | 1.23 (1.12-1.35) | 1.31 (1.14-1.51) | 1.20 (1.09-1.32) | 1.17 (1.05-1.30) |
| **Other immunosuppressive condition** | 1.69 (1.21-2.34) | 2.01 (1.25-3.25) | 1.52 (1.06-2.19) | 1.66 (1.16-2.39) |

FOOTNOTES: Models adjusted for age using a 4-knot cubic spline age spline, except for estimation of age group effects. \*Ethnicity effect estimated from a model restricted to those with recorded ethnicity. \*<sup>2</sup>OCS = oral corticosteroids. Recent OCS use defined as in the year before baseline. \*<sup>3</sup>HbA1c classification based on latest

